# Oral potassium supplementation lowers clinic blood pressure in untreated primary hypertension—systematic review and meta-analysis of randomized trials

**DOI:** 10.64898/2025.12.08.25341863

**Authors:** Javier Ravichagua Ashiyama, Felix Medina Palomino, Cesar Loza Munarriz, Julio Poterico Rojas, Shirley Ramirez Sanchez

## Abstract

**Background:** Potassium has biologic plausibility to lower blood pressure (BP) via natriuresis, vascular effects, and RAAS modulation. The clinical effect in untreated primary hypertension remains uncertain.

**Objectives:** To estimate the effect of oral potassium supplementation on clinic and 24-h ambulatory BP (ABPM) and to summarize safety.

**Methods:** We included randomized trials (parallel or crossover) in adults (≥18 y) with untreated primary hypertension comparing oral potassium supplements vs placebo, sodium chloride, or no treatment. Trials without end-of-treatment BP or without performed 24-h urinary potassium measurements were excluded. Sources: PubMed, Embase, CENTRAL, Scopus, CINAHL, trial registries (through Jan 31, 2025; updated Oct 1, 2025). Random-effects meta-analyses (REML, Knapp–Hartung) pooled end-of-treatment mean differences (mmHg) and risk ratios (RR); prediction intervals (PI) were reported. Certainty was appraised with GRADE.

**Results:** Nineteen trials (803 participants; 14 crossover, 5 parallel; median duration 4 weeks) met criteria. Potassium lowered clinic BP: SBP −7.2 mmHg (95% CI −11.1 to −3.4; PI−24.5 to 10.1; I² = 92.3%) and DBP −4.0 mmHg (−6.2 to −1.7; PI −14.1 to 6.1; I² = 94.8%). ABPM effects were smaller and imprecise: SBP −2.4 mmHg (−5.2 to 0.4; I² = 55.0%) and DBP −1.6 mmHg (−4.2 to 1.1; I² = 84.3%). Any adverse event was more frequent (RR 2.41; ≈40 more per 1000, 17–75), with small absolute increases in total withdrawals (RR 1.32; ≈4 more per 1000, 0–9) and AE-related withdrawals (RR 1.28; 0–10 more per 1000). Signals suggested larger benefits with higher baseline BP, longer duration, and in parallel-group trials.

**Discussion:** Our findings reinforce the physiological plausibility that potassium exerts antihypertensive effects primarily through natriuresis and vascular modulation, rather than a simple dose-dependent mechanism. The modest mean effect, together with wide prediction intervals, underscores the role of individual sodium–potassium balance, baseline BP, and intervention duration in determining response. These data support a shift from universal supplementation toward context-specific implementation, guided by biochemical monitoring and patient characteristics.

**Conclusions:** In untreated primary hypertension, oral potassium supplementation yields modest, clinically relevant reductions in clinic BP, while ABPM effects are smaller and very uncertain. Given mostly mild AEs and small absolute increases in withdrawals, supplementation may be conditionally considered alongside sodium reduction and routine laboratory monitoring. Adequately powered parallel trials (≥8–12 weeks) with prespecified ABPM and standardized safety reporting are needed.

## INTRODUCTION

### 3. Rationale

Hypertension is a leading, preventable driver of cardiovascular morbidity and mortality. In parallel, low potassium intake is common in many settings. Oral potassium supplementation has strong biologic plausibility to lower blood pressure through natriuresis and modulation of vascular tone and the renin–angiotensin–aldosterone system. Yet the clinical evidence in adults with untreated primary hypertension remains unsettled.

Randomized trials are dispersed, frequently small and of short duration, and often use crossover designs that may be vulnerable to carryover if washout is insufficient. Reporting of safety, adherence, and background sodium intake is inconsistent, and relatively few studies assess ambulatory blood pressure monitoring (ABPM), which is less affected by white-coat phenomena and better reflects out-of-office control. Prior summaries mix dietary interventions with pharmacologic supplementation, include treated populations, or provide limited synthesis of ABPM and adverse outcomes, leaving clinicians without a clear, decision-oriented estimate of effect for the specific scenario of untreated primary hypertension.

Given the potential for a low-cost, scalable intervention—balanced against concerns about hyperkalemia in susceptible patients—a focused, up-to-date synthesis is needed to inform practice and guideline development. Our objective was to provide a comprehensive, transparent summary of randomized evidence on oral potassium supplementation in adults with untreated primary hypertension, estimating its effect on clinic and ambulatory blood pressure at the end of treatment and summarizing withdrawals and adverse events, while clarifying the main sources of uncertainty that affect applicability in clinical care.

### 4. Objectives

The primary objective was to determine, among adults (≥18 years) with untreated primary hypertension, the effect of oral potassium supplementation, compared with placebo, sodium chloride or no treatment; on clinic and 24-h ambulatory systolic and diastolic blood pressure at end of treatment (mmHg). Secondary objectives were to summarize withdrawals, withdrawals due to adverse events and any adverse events. Exploratory analyses examined dose–response and between-study heterogeneity using achieved urinary potassium as an exposure/adherence indicator, not as an outcome.

## METHODS

### 5. Eligibility criteria

Randomized clinical trials, with either parallel or crossover design, were included if they evaluated oral potassium supplementation compared with placebo, sodium chloride or no intervention, in adults (≥18 years) with untreated primary hypertension. Studies were excluded if they involved secondary hypertension, pregnancy or co-supplementation with other minerals (e.g., magnesium or calcium), as well as those based solely on potassium-enriched salt substitutes or dietary modifications. No restrictions were applied regarding year, language or publication status; published articles, unpublished manuscripts, and conference abstracts were all considered. Studies were excluded if end-of-treatment blood pressure was not measured, if 24-hour urinary potassium measurements were not performed (to confirm exposure to the assigned potassium supplementation and adherence), or if essential data could not be extracted or derived. For synthesis, studies were grouped by comparator type (placebo/no intervention vs sodium chloride), potassium supplementation form and trial design (parallel vs crossover). These restrictions were applied to ensure comparability across interventions and accurate quantification of supplemented potassium doses.

### 6. Information sources

We systematically searched PubMed, Ovid Embase, the Cochrane Central Register of Controlled Trials (CENTRAL), Scopus and CINAHL (via EBSCOhost) from database inception to January 31, 2025. Searches were updated before data synthesis. In addition, we screened trial registries including ClinicalTrials.gov and the World Health Organization International Clinical Trials Registry Platform (WHO ICTRP), to identify ongoing or unpublished studies. The reference lists of all included trials and relevant systematic reviews were also examined to capture additional records. When essential data were missing, study authors were contacted directly for clarification or to obtain unpublished information.

### 7. Search strategy

The search strategy was developed using a PICO-based framework and adapted for each database. The PubMed search was drafted first and then translated for Ovid Embase, CENTRAL, Scopus and CINAHL using database-specific subject headings and syntax rules. The strategy combined controlled vocabulary (e.g., MeSH terms) and free-text terms for *potassium supplementation*, *blood pressure* and *hypertension*, limited to human adults without language or publication date restrictions.

A sample PubMed search string was:

(“potassium”[MeSH Terms] OR “potassium supplementation”[tiab] OR “oral potassium”[tiab]) AND (“blood pressure”[MeSH Terms] OR “blood pressure”[tiab] OR “hypertension”[tiab]) AND (“randomized controlled trial”[Publication Type] OR randomized[tiab] OR randomised[tiab] OR RCT[tiab]) AND (“adult”[MeSH Terms] OR adult[tiab]) NOT (animals[MeSH Terms] NOT humans[MeSH Terms])

Searches covered database inception to January 31, 2025, with an update performed in October 1^st^, 2025 (immediately before data synthesis). Equivalent strategies were implemented in other databases, adapting controlled vocabulary (e.g., Emtree in Embase) and syntax as needed. No language filters were applied.

The draft strategy was peer-reviewed by an information specialist following the Peer Review of Electronic Search Strategies (PRESS) checklist. Full, line-by-line search strategies for all databases, including Boolean operators and field tags, are provided in the **Supplementary Materials**.

### 8. Selection process

All records were de-duplicated in Zotero and screened in two stages (titles/abstracts, then full texts) by two independent reviewers (JRA, SRS). Disagreements at either stage were resolved by discussion or, when needed, adjudication by a third reviewer (JPR). One study required translation; its full text was translated using an automated tool and then screened accordingly. ASReview LAB v2.0.2 was used exclusively to prioritize the screening order and not to make inclusion/exclusion decisions. The software’s seed-selection options were used, and to mitigate false negatives we manually reviewed every abstract up to 200 records after the last included study, which amounted to approximately 20% of the database. Reasons for full-text exclusion were recorded for the PRISMA flow diagram. No records were eliminated by automation tools.

### 9. Data collection process

Two reviewers (JRA, SRS) independently and in duplicate extracted data from each eligible report using a piloted Excel workbook with predefined fields and validation checks; discrepancies were resolved by consensus or, when needed, adjudication by a third reviewer (JPR). One non-English study required translation and an automated translation tool was used both to determine eligibility and to enable data extraction, translated passages were reviewed by the extractors. No automation tools were used to extract data from reports and no software was used to digitize values from figures; only numerically reported tables or text were extracted. When multiple reports corresponded to the same study, we applied a prespecified decision rule: prioritize the report with the most complete blood-pressure data and cross-check against companion reports to resolve inconsistencies. Attempts to contact study investigators to obtain or confirm missing information were unsuccessful, in some cases due to the age of the trials and lack of contact details, for this, extractions relied on available data only.

### 10a. Data items (outcomes)

The prespecified primary outcomes were clinic systolic and diastolic blood pressure (SBP, DBP) at end of treatment (mmHg). Ambulatory blood pressure (24-h SBP/DBP by ABPM) was treated as a co-primary outcome. Secondary outcomes were total withdrawals, withdrawals due to adverse events and any adverse event. Urinary potassium (24-h or aliquot) was collected solely as an exposure/adherence indicator (for dose–response analyses) and not as an outcome.

We sought all results compatible with each outcome domain at the end-of-treatment time point. When multiple measures were reported for the same domain, we applied a prespecified hierarchy: for clinic BP, supine was prioritized over seated and seated over standing; for ambulatory BP, 24-h averages were prioritized over daytime and then nighttime. If several analytic summaries existed for the same outcome (e.g., multiple time windows or repeated assessments), we extracted the measure corresponding to the final end-of-treatment assessment. No post-hoc changes were made to the inclusion or definitions of outcome domains or to their relative importance.

### 10b. Data items (other variables)

Beyond outcomes, we extracted study- and arm-level characteristics: country and year, trial design (parallel or crossover), sample sizes per arm, participant characteristics (mean age, sex), baseline clinic/ambulatory BP, intervention details (potassium salt form and assigned dose, mmol/day) and duration (weeks), any sodium-intake modification, adherence/exposure markers including achieved urinary potassium and sodium (24-h or aliquot), blood-pressure measurement posture (supine, seated, standing) and ambulatory summary (24-h, daytime, nighttime), withdrawals (total and due to adverse events) and any adverse event, and trial registration identifiers where reported. Selection of data items was informed by the Cochrane Handbook for Systematic Reviews of Interventions (v6.3). When only SEs, CIs, p-values or medians/IQRs were reported, we converted to means/SDs using standard Cochrane transformations. For crossover trials lacking within-person variance we imputed a within-person correlation of r = 0.5. When BP was reported by sex, pooled means/SDs were computed weighting by subgroup sizes. If essential information remained unavailable, extractions relied on the reported data and reasons for missingness were documented.

### 11. Study risk of bias assessment

Risk of bias was assessed with the Cochrane Risk of Bias 2 (RoB 2) tool at the outcome level for SBP and DBP. Two reviewers (JRA, SRS) conducted independent assessments; disagreements were resolved by discussion or adjudication by a third reviewer (JPR). The five RoB 2 domains were evaluated (randomization process, deviations from intended interventions, missing outcome data, measurement of the outcome, selection of the reported result) and, overall judgments were derived according to the RoB 2 decision rules. No adaptations of the tool and no automation were used. Risk-of-bias results were tabulated and informed sensitivity analyses (excluding high risk-of-bias studies) and exploratory stratified analyses.

### 12. Effect measures

For continuous outcomes (clinic and ambulatory SBP/DBP at end of treatment), effects were synthesized and presented as mean differences (mmHg) on the original scale; by convention, negative values favor potassium. In crossover trials, paired mean differences (or their standard errors) were used when available; otherwise, standard errors were derived as specified in the Methods. For binary outcomes (total withdrawals, withdrawals due to adverse events and any adverse event), effects were summarized as risk ratios (RRs) with 95% confidence intervals. We did not pool change-from-baseline values, only end-of-treatment measurements were synthesized. For zero-event trials, we applied a continuity correction of 0.5 to all four cells and increased arm totals accordingly; studies with zero events in both arms were retained after correction. Risk ratios were then pooled using a Mantel–Haenszel random-effects model with Hartung–Knapp adjustment. When a study included more than one eligible potassium arm versus a single control, we treated each comparison as a separate study-level effect while accounting for the shared control. For continuous outcomes (mean differences), we split the control group equally across comparisons, which is equivalent to inflating the control standard error by √K (K = number of comparisons). For binary outcomes, we split events and totals in the shared control across comparisons.

### 13a. Synthesis methods (eligibility for synthesis)

For each planned synthesis we included randomized controlled trials that matched the protocol PICO and that reported enough information to compute effect estimates for the relevant outcome. For continuous outcomes (systolic and diastolic blood pressure at end of follow-up), studies were eligible if they provided arm-level means with a measure of precision or a mean difference with its standard error. For harms, three binary outcomes were analyzed at the participant level: withdrawal due to adverse events, total withdrawals, and any adverse event. Studies without sufficient data for a given synthesis were not pooled for that synthesis.

### 13b. Synthesis methods (preparing for synthesis)

We harmonized units (mmHg for blood pressure). When only standard deviations were reported, we converted SD to SE using SE = SD/√n. For cross-over designs that reported arm-level summaries, we computed the standard error of the treatment contrast using a fixed within-person correlation of 0.5. Duplicate reports were resolved by prioritizing the report with the most complete data.

### 13c. Synthesis methods (tabulation and graphical methods)

We presented structured evidence tables of study characteristics and outcomes and we produced forest plots for each meta-analysis. For continuous outcomes we displayed forests that include, per study, arm sizes, means and SDs alongside the study effect and weight; for harms we produced separate forest plots for risk ratio and risk difference. Additional figures included a benefit–risk scatterplot (blood pressure effect vs absolute difference in AE withdrawals), dose–response curves using restricted cubic splines (1-stage approach), and probability curves showing the model-based probability that the blood-pressure reduction is at least 5 or 10 mmHg across exposure levels.

### 13d. Synthesis methods (statistical synthesis methods)

Meta-analyses of continuous outcomes used inverse-variance random-effects models with restricted maximum likelihood (REML) for τ² and Hartung–Knapp adjustments for summary-effect confidence intervals; we reported τ², I² and prediction intervals (PIs). Binary harms (AE withdrawals, total withdrawals, any AE) were synthesized with random-effects Mantel–Haenszel models for risk ratio and risk difference. All analyses were conducted in R 4.5.1 using packages: metafor, meta, tidyverse, janitor, ggplot2, ggrepel, ragg, patchwork, writexl and splines.

### 13e. Synthesis methods (methods to explore heterogeneity)

When studies reported clinic and ambulatory BP at the same time point in the same participants, we had planned to fit a multivariate random-effects model (REML) assuming a within-study correlation of 0.5 between outcomes. As pre-specified, multivariate pooling would only be performed if at least 8 studies contributed both outcomes to ensure model stability. Because only 6 studies had the required paired data, we did not implement the multivariate model and instead synthesized clinic and ambulatory BP separately using univariate random-effects models with Knapp–Hartung inference. We reported τ², I² and PIs. Pre-specified univariable meta-regressions were performed on study-level moderators defined in the protocol (comparator type, clinical posture, salt form, study design, and overall risk of bias). Dose–response analyses were performed using 1-stage restricted cubic splines for both assigned dose and achieved ΔK. Knots were placed at the 10th, 50^th^ and 90th percentiles of the observed exposure, and we reported spline predictions with CIs along the observed range. As a sensitivity analysis, spline models were re-estimated with four knots at the 5th/35th/65th/95th percentiles. Post hoc exploratory analyses included baseline urinary sodium and potassium, achieved urinary K:Na ratio and a multivariable meta-regression to adjust for key covariates: for SBP, duration, baseline SBP (centered) and achieved urinary K:Na ratio; for DBP, duration, baseline DBP (centered), achieved urinary K:Na ratio and baseline urinary sodium (centered).

### 13f. Synthesis methods (sensitivity analyses)

Sensitivity analyses focused on data handling and modeling choices that could plausibly influence the pooled effects. For AE withdrawals, we verified consistency across effect measures by synthesizing both risk ratio and risk difference under the same random-effects framework and continuity-correction rule. The planned first-period-only re-analysis for crossover trials could not be performed because included reports did not provide the necessary period-specific summaries. Post-hoc, we applied SIMEX to assess measurement-error impact on meta-regressions using Δ urinary K.

### 14. Reporting bias assessment

We assessed the risk of bias due to missing results (reporting biases) for each meta-analysis meeting the conventional threshold of ≥10 studies. Specifically, we examined funnel plots and interpreted any asymmetry qualitatively while considering between-study heterogeneity. We performed Egger’s regression under a random-effects framework with a two-sided α of 0.10 given limited power and applied Duval and Tweedie’s trim-and-fill only as a sensitivity analysis rather than to replace primary estimates. When fewer than 10 studies were available, we did not run formal small-study tests and provided a narrative assessment. In addition, we used the ROB-ME (Risk Of Bias due to Missing Evidence) tool to generate domain-level and overall judgments for the meta-analyses of primary and key secondary outcomes. Assessments were carried out by two reviewers. Where possible, we compared registered protocols or registry entries and methods sections against the reported outcomes to evaluate selective non-reporting; some registrations were available and reviewed.

### 15. Certainty assessment

We assessed the certainty of evidence using the GRADE approach for all key outcomes (SBP, DBP, withdrawals due to adverse events, total withdrawals, and any adverse event). Randomized evidence started at high certainty and was rated down, when applicable, for risk of bias (informed by RoB 2 judgments), inconsistency (direction and magnitude of effects together with I², τ², and PIs), indirectness (population, intervention, comparator, outcomes), imprecision (judged against the null) and reporting bias. Two reviewers conducted GRADE independently and resolved differences by discussion (with adjudication when needed). We compiled Summary of Findings tables and documented explicit footnoted rationales for each downgrade; no automation tools were used.

## RESULTS

### 16. Study selection

In the study selection, the PRISMA flow diagram (Figure 1) summarizes identification, screening and inclusion. After deduplication and automation-assisted exclusions, 1294 records were screened, 35 full-text reports were assessed and 19 studies were included. Reports identified by other methods (n = 32) were assessed and all were duplicates of database records. Some examples of full-text reports that might appear eligible but were excluded, with the main reason, are: Svetkey (1986), no urinary potassium measurements; He (2005), two active interventions without a control arm; Smith (1983), nonrandomized study without a concurrent control group.

**Figure 1.**
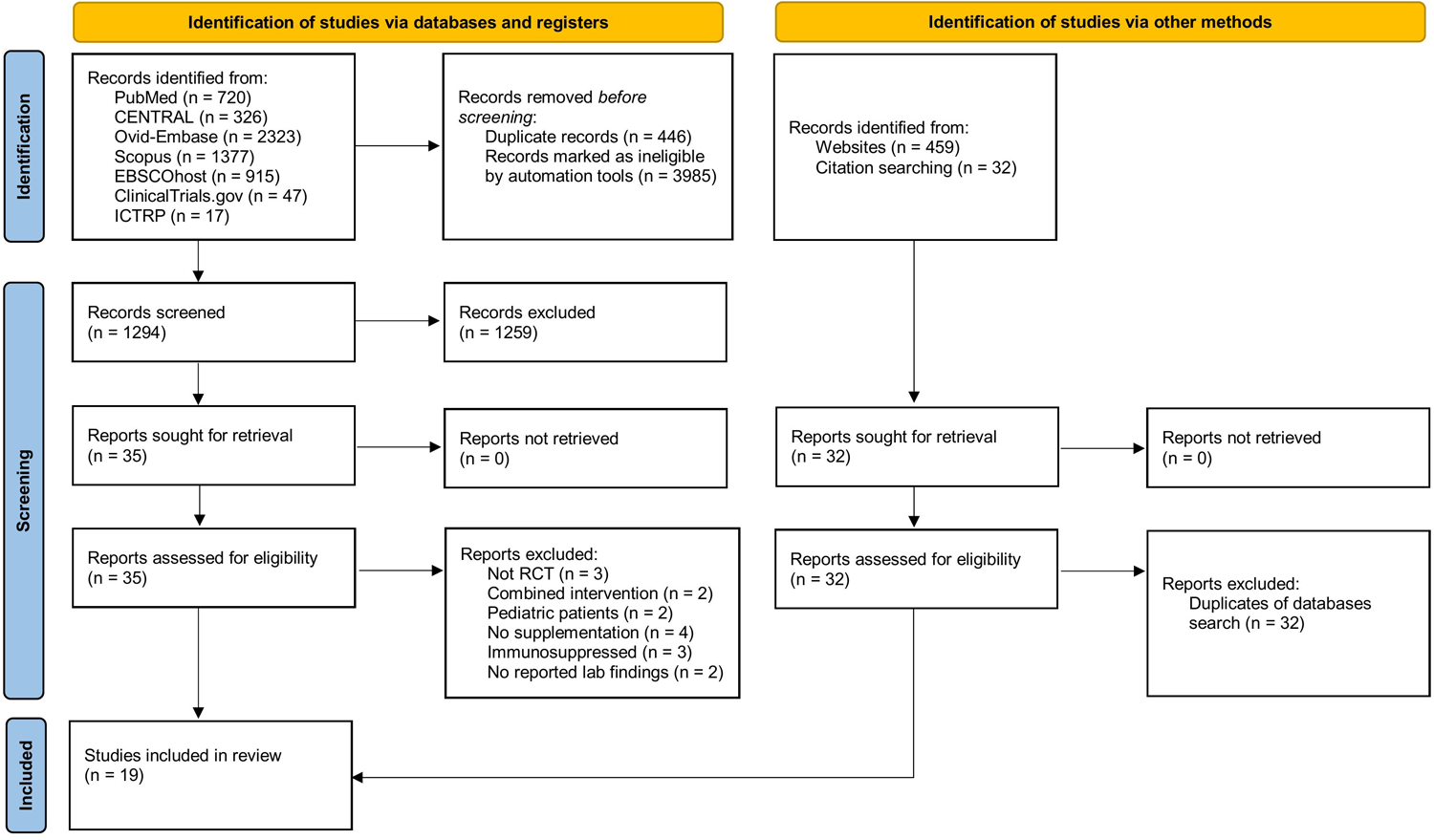
PRISMA 2020 flow diagram of the study selection process.

### 17. Study characteristics

We included 19 randomized trials (1–19) in adults with untreated primary hypertension (**Table 1**). The total sample comprised 803 participants. Trial designs were predominantly crossover (14 of 19) with the remainder parallel. Studies were conducted between 1982 and 2021 across the United Kingdom, Italy, Netherlands, China, South Africa, Kenya, Germany, India, New Zealand, United States and Chile. Per-trial sample sizes ranged from 12 to 150 participants. Most trials enrolled both sexes (18/19), one enrolled women only. Where reported, mean ages spanned approximately 18 to 79 years. Reported female counts per trial ranged from 4 to 90, summing to 385 across trials with available counts.

**Table 1.**
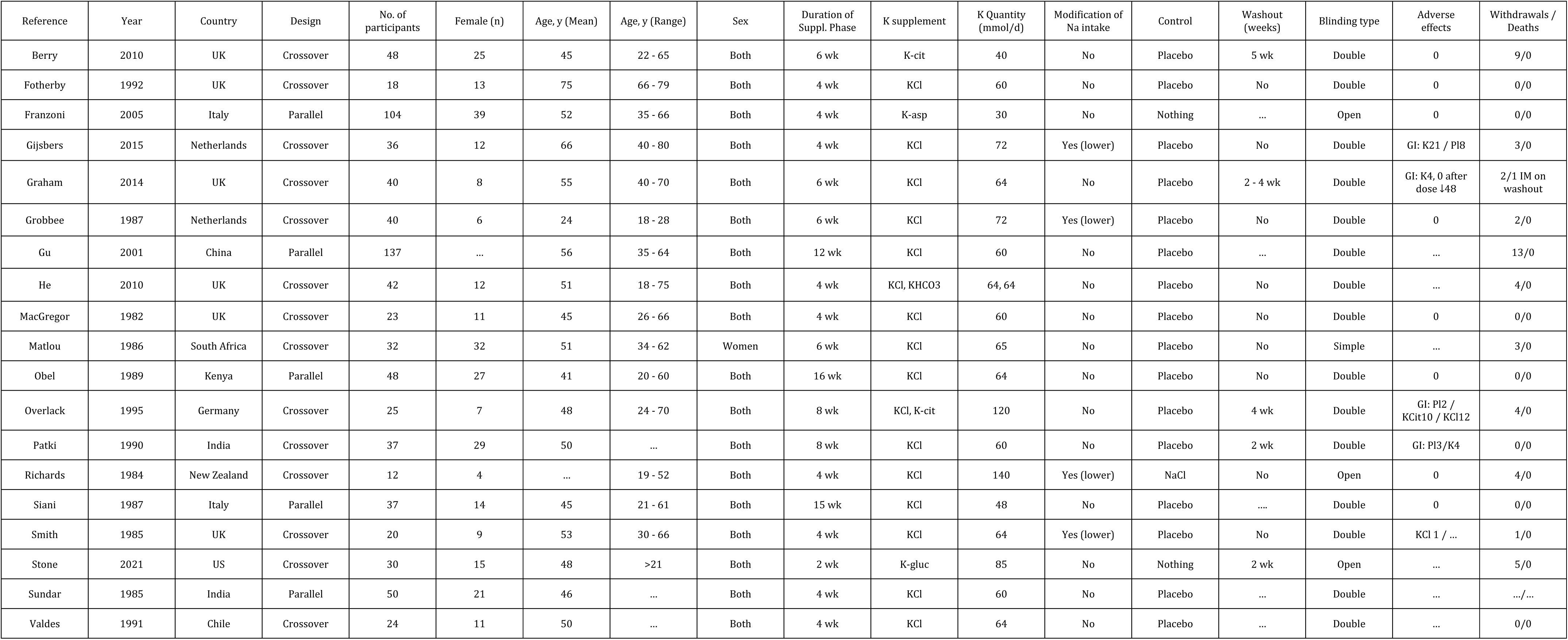
Key characteristics of included studies. KCl, potassium chloride; K-cit, potassium citrate; K-asp, potassium aspartate; KHCO3, potassium bicarbonate; K-gluc, potassium gluconate; NaCl, sodium chloride; wk, weeks.

Potassium was most often given as potassium chloride, with less frequent use of citrate, aspartate, bicarbonate or gluconate. Assigned doses across intervention arms typically ranged from 30 to 140 mmol/day (median 64; IQR 60–66). Supplementation phases were short to moderate in length, most commonly 4 weeks (median 4; range 2–16). Comparators were predominantly placebo (16/19), with two no-treatment controls and one sodium chloride control. A reduced sodium intake was co-implemented in four trials.

End-of-treatment clinic blood pressure was the primary outcome across trials, six trials additionally reported ambulatory blood pressure. Adherence/exposure markers such as urinary potassium and where available urinary sodium, were collected to contextualize exposure. Blinding was most often double-blind, with a minority open-label or single-blind. Five crossover trials reported washout periods (typically 2–5 weeks), although the others either did not implement a washout or did not report one.

### 18. Risk of bias in studies

We assessed risk of bias using RoB 2 at the outcome level for clinic systolic and diastolic blood pressure (SBP, DBP) and for 24-h ambulatory SBP and DBP. Two reviewers worked independently with adjudication. Domain-level judgments (D1–D5) are provided in **Supplementary Tables S1–S7**. In addition to the outcome-level assessments, we applied RoB 2 at the study level to derive an overall rating for each trial (low risk, some concerns, or high risk). These study-level ratings were used in the pre-specified subgroup analyses by overall risk of bias, and the corresponding table with domain-level rationales is provided in the **Supplementary Table S8**.

For clinic BP (SBP and DBP), overall judgments were most often “some concerns.” The main reasons were D3, because several trials analyzed completers or had modest post-randomization loss that was not imputed—an issue that is more prominent in cross-over designs and can bias end-of-period clinic means if attrition relates to outcome—and D4, where a subset relied on manual auscultatory clinic BP without explicit assessor blinding or random-zero/automatic devices, increasing the possibility of detection bias. Trials that used automated or random-zero devices, applied standardized rest with multiple readings and had minimal balanced attrition, were typically judged low risk for the clinic outcomes. D5 was generally low risk because clinic endpoints and analysis windows were prespecified and consistently reported.

For 24-h ABPM (SBP and DBP), judgments were more favorable. Almost all assessments were low risk for D4, reflecting automated devices (e.g., Spacelabs), predefined day and night windows, and minimum-reading criteria. Residual “some concerns” arose mainly from D3 when ABPM datasets were incomplete or analyzed in completers. D5 was low risk across ABPM analyses because prespecified windows (24-h, daytime, nighttime) were reported without selective emphasis.

For safety outcomes—total withdrawals, withdrawals due to adverse events and any adverse event—domain-level judgments for contributing trials were low risk in D3, D4 and D5. Most reports did not detail the adverse-event ascertainment schedule and frequently relied on participant self-report; however, the trials included in our meta-analyses provided arm-level counts with clear denominators and comparable follow-up under blinded conditions, and withdrawals are objective outcomes. Reporting covered the prespecified safety endpoints rather than selected subcategories. Cross-over trials without arm-level denominators or without a comparable capture window were not meta-analyzed and therefore were not assessed for within-study risk of bias for these outcomes.

### 19. Results of individual studies

Nineteen randomized trials contributed 21 comparisons, two multi-arm trials contributed two comparisons each. Across the 19 trials, potassium supplementation reduced clinic blood pressure at end of treatment. The pooled effect for clinic SBP was −7.2 mmHg (95% CI −11.1 to −3.4; PI −24.5 to 10.1; I² = 92.3%), and for clinic DBP was −4.0 mmHg (95% CI −6.2 to −1.7; PI −14.1 to 6.1; I² = 94.8%). Effects generally favored supplementation, with substantial between-study heterogeneity.

Among studies reporting ambulatory (24-h) blood pressure, six trials contributing seven comparisons showed small and imprecise reductions. The pooled mean difference for SBP was −2.4 mmHg (95% CI −5.2 to 0.4; PI −8.8 to 3.9; I² = 55.0%), and for DBP was −1.6 mmHg (95% CI −4.2 to 1.1; PI −8.8 to 5.7; I² = 84.3%). Effects tended to favor supplementation, but overall tests were not statistically significant. Individual study results for clinic SBP and DBP are shown in Figures 2-3.

**Figure 2.**
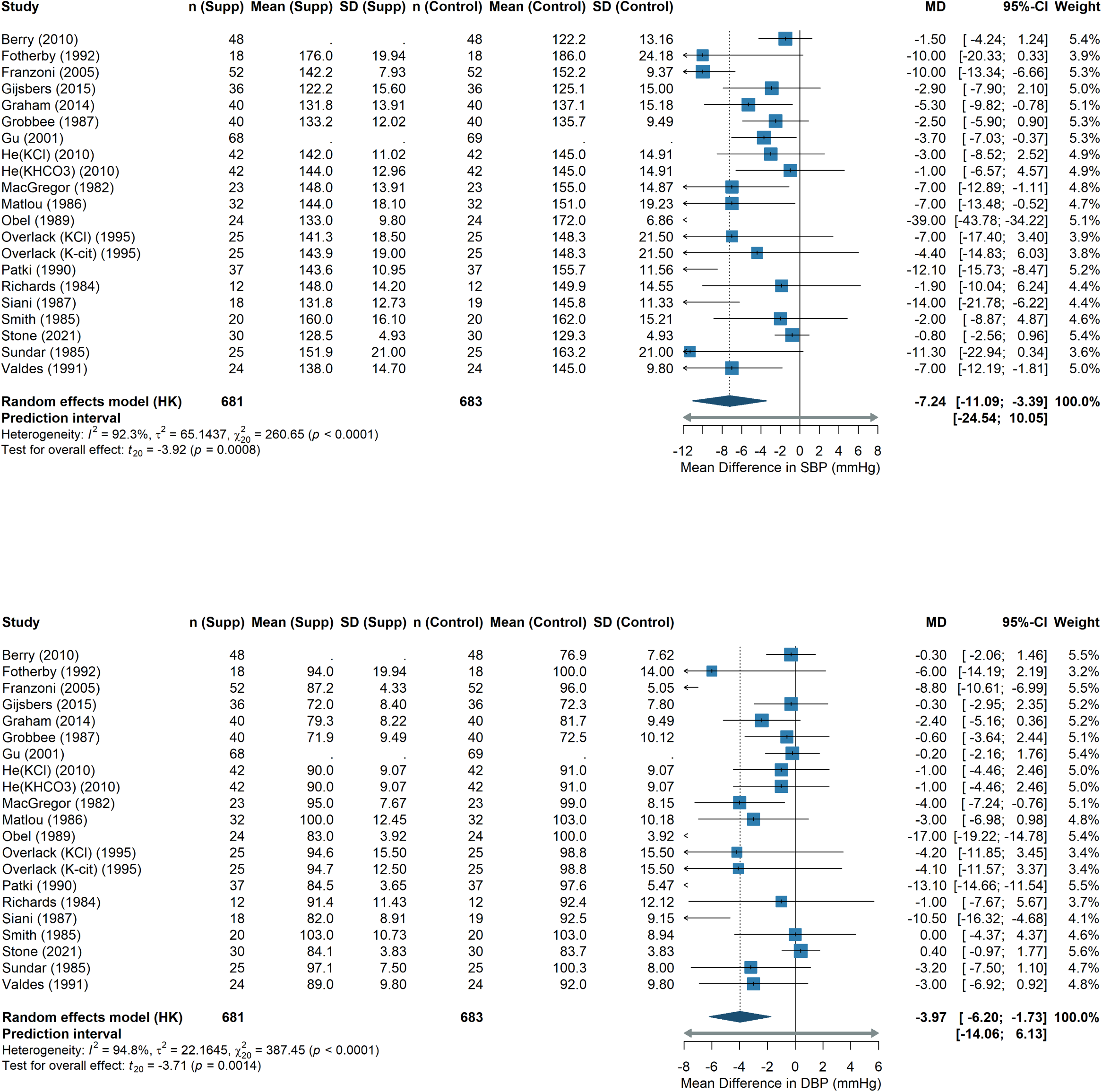
Forest plots of end-of-treatment blood pressure: SBP (top) and DBP (bottom).

**Figure 3.**
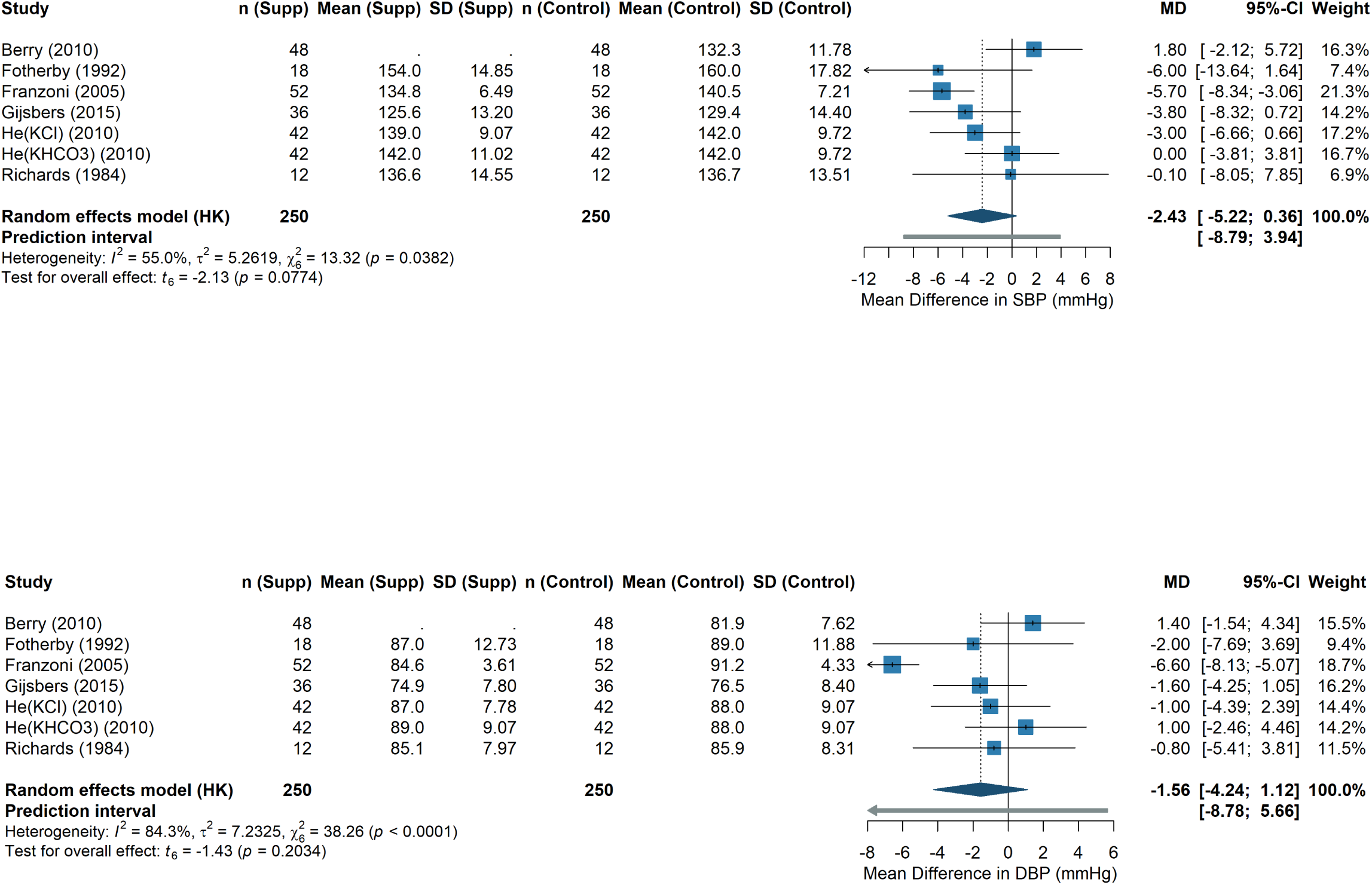
Ambulatory blood pressure at end of treatment: SBP (top) and DBP (bottom).

### 20a. Results of syntheses (characteristics of contributing studies)

#### Clinical synthesis (office BP: SBP/DBP at end of treatment)

Contributing studies were randomized trials in adults with untreated primary hypertension, mostly crossover designs with several parallel trials. Interventions were oral potassium supplements expressed in mmol/day, with potassium chloride as the predominant salt and less frequent use of citrate, bicarbonate, aspartate or gluconate; comparators were mainly placebo or no intervention and, in one case, sodium chloride. Office blood pressure was extracted following the pre-specified hierarchy for clinical measurements (priority supine, then seated and standing) and synthesized as mean difference at end of treatment. Risk of bias was most often judged ‘some concerns’ mainly due to D3 (use of completer analyses or modest post-randomisation loss) and D4 (manual auscultatory measurement without explicit assessor blinding). D5 was generally low risk because endpoints and analysis windows were prespecified and consistently reported.

#### Ambulatory synthesis (24-hour ABPM)

This synthesis included trials reporting 24-hour SBP/DBP averages at end of treatment. This subset shares the same population framework, salt forms and comparators as the clinical synthesis; and was analyzed as a co-primary outcome. When paired clinical/ambulatory data were available within the same study, joint modeling assuming within-study correlation was contemplated, otherwise univariate syntheses were performed. The risk-of-bias profile was more favorable than for clinic BP: D4-D5 were low risk in all ABPM assessments owing to automated devices with predefined day/night analysis windows, and remaining ‘some concerns’ arose mainly from D3. The contributing studies and details appear in the corresponding figures and the characteristics table.

#### Safety synthesis (total withdrawals, withdrawals due to adverse events, and any adverse event)

This synthesis included trials that reported withdrawals and/or adverse events under the same comparators and durations as the efficacy syntheses. We meta-analyzed withdrawals due to adverse events reporting risk ratios as the primary measure and risk differences as complementary absolute effects. For total withdrawals and for any adverse event, we synthesized risk ratios only. Risk of bias was assessed for all three safety outcomes. For trials contributing to the meta-analyses, D3, D4 and D5 were judged low risk because studies reported arm-level counts with clear denominators and comparable follow-up under blinded protocols, and reporting covered the prespecified safety endpoints.

### 20b. Results of syntheses (results of statistical syntheses)

#### Clinic BP (end of treatment)

Across 21 comparisons from 19 trials, potassium supplementation lowered office blood pressure. The pooled mean difference for SBP was −7.2 mmHg (95% CI −11.1 to −3.4; PI −24.5 to 10.1; I² = 92.3%) and for DBP was −4.0 mmHg (95% CI −6.2 to −1.7; PI −14.1 to 6.1; I² = 94.8%). Negative values favor potassium. Forest plots are shown in **Figure 2**.

#### Ambulatory BP (24-h ABPM, end of treatment)

In six trials contributing seven comparisons, pooled effects suggested small and imprecise reductions: SBP −2.4 mmHg (95% CI −5.2 to 0.4; PI −8.8 to 3.9; I² = 55.0%) and DBP −1.6 mmHg (95% CI −4.2 to 1.1; PI −8.8 to 5.7; I² = 84.3%). Overall tests were not statistically significant. See **Figure 3**.

#### Safety (AE withdrawals)

A random-effects synthesis showed a small increase in relative risk (RR 1.3; 95% CI 1.0–1.6; I² = 0%; k = 17; **Supplementary Figure S1**). In absolute terms, the pooled risk difference was 0.0 per 1000 participants (95% CI 0 to 1; I² = 0%; **Supplementary Figure S2**), consistent with a near-zero absolute change.

#### Safety (any adverse event)

Thirteen trials contributed data; potassium increased the risk of experiencing at least one adverse event (RR 2.41, 95% CI 1.63–3.56; I² = 0%; see **Supplementary Figure S3**).

#### Safety (total withdrawals)

Twelve trials showed a modest increase in overall withdrawals (RR 1.32, 95% CI 1.03–1.70; I² = 0%; **Supplementary Figure S4**).

To complement the safety synthesis, we generated a by-study benefit–risk scatterplot (**Supplementary Figure S5**) plotting the end-of-treatment mean difference in systolic blood pressure (mmHg; negative values favor potassium) on the x-axis against the absolute risk difference in withdrawals due to adverse events per 100 participants on the y-axis. Most trials clustered near zero absolute change in AE-related withdrawals while showing SBP reductions, whereas a small subset (e.g., Overlack 1995, Gijsbers 2015, Smith 1985) displayed higher AE-related withdrawals with limited blood-pressure benefit.

### 20c. Results of syntheses (investigations of heterogeneity)

#### A. Subgroup analyses

##### Comparator type

For systolic blood pressure (SBP), trials using placebo or no treatment as control (k = 20) yielded a pooled mean difference (MD) of −7.49 mmHg (95% CI −11.50 to −3.48), whereas the single NaCl-controlled trial (k = 1) showed a non-significant MD of −1.90 mmHg (95% CI −10.04 to 6.24). For diastolic blood pressure (DBP), the placebo/no-treatment subgroup (k = 20) produced an MD of −4.08 mmHg (95% CI −6.40 to −1.76); NaCl-controlled MD of −1.00 mmHg (95% CI −7.67 to 5.67). Given the extreme imbalance between subgroups (20 vs 1 study), these findings should not be interpreted as evidence that the choice of comparator modifies the effect.

##### Potassium supplementation form

For SBP, trials using potassium chloride (KCl) supplements (k = 16) showed a pooled MD of −8.48 mmHg (95% CI −13.39 to −3.56), whereas studies using other potassium salts (k = 5) yielded a non-significant MD of −3.44 mmHg (95% CI −8.58 to 1.69). For DBP, the corresponding pooled estimates were −4.39 mmHg (95% CI −7.16 to −1.61) for KCl and −2.65 mmHg (95% CI −7.60 to 2.30) for other potassium salts. Only KCl reached nominal significance in subgroup pooling, but meta-regression did not support salt form as an effect modifier.

##### Study design

Parallel-group trials (k = 5) showed a larger mean reduction in SBP of −15.68 mmHg (95% CI −32.84 to 1.49) compared with crossover studies (k = 16; −4.34 mmHg, 95% CI −6.21 to −2.47). For DBP, parallel trials (k = 5) yielded a pooled mean difference of −7.93 mmHg (95% CI −16.18 to 0.33), whereas crossover designs (k = 16) produced −2.64 mmHg (95% CI −4.57 to −0.71). Although confidence intervals were wide in the smaller parallel subset, the direction of the effect was consistent, with crossover studies tending to show smaller reductions in blood pressure.

##### Participant posture at measurement

Random-effects meta-analyses stratified by posture showed, for SBP, pooled MDs of −8.93 mmHg (95% CI −15.09 to −2.78; I² = 94.3%) in supine measurements, −4.27 mmHg (95% CI −7.13 to −1.41; I² = 74.1%) in seated measurements, and −11.14 mmHg (95% CI −21.15 to −1.13; I² = 95.2%) in standing measurements (test for subgroup differences: χ² = 4.27, df = 2, p = 0.118). For DBP, pooled MDs were −4.77 mmHg (95% CI −8.18 to −1.37; I² = 95.6%) supine, −2.59 mmHg (95% CI −5.29 to 0.11; I² = 90.8%) seated, and −6.14 mmHg (95% CI −11.63 to −0.65; I² = 94.7%) standing, with no significant subgroup differences (χ² = 2.52, df = 2, p = 0.283). Although point estimates tended to be more negative in standing measurements, posture did not materially modify the effect.

##### Overall risk of bias

Only two trials were judged at low risk of bias and 17 were rated as some/high risk. For SBP, low-risk trials showed a modest and imprecise reduction (−4.22 mmHg; 95% CI −19.39 to 10.95; I² = 0%), whereas some/high-risk trials showed a larger and statistically significant decrease (−7.60 mmHg; 95% CI −11.86 to −3.33; I² = 93.1%); the subgroup difference was not statistically significant (p = 0.15). For DBP, pooled effects were −1.31 mmHg (95% CI −14.64 to 12.02; I² = 13.4%) in low-risk trials and −4.27 mmHg (95% CI −6.72 to −1.82; I² = 95.2%) in some/high-risk trials, with a borderline subgroup difference (p = 0.05). Given the very small number of low-risk studies, these contrasts should be interpreted cautiously.

#### B. Meta-regression

##### Categorical moderators (REML with Hartung–Knapp; k = 21)

Comparator type was not associated with differential effects (SBP β = 5.6 mmHg, 95% CI −13.6 to 24.8, p = 0.55; DBP β = 3.1 mmHg, 95% CI −8.9 to 15.1, p = 0.60). Potassium salt form was likewise non-significant (SBP β = 5.0 mmHg, 95% CI −3.9 to 13.8, p = 0.26; DBP β = 1.7 mmHg, 95% CI −3.5 to 7.0, p = 0.50). Study design did influence results: relative to parallel trials, crossover studies showed smaller mean differences (SBP β = 11.2 mmHg, 95% CI 3.5 to 18.9, p = 0.007; DBP β = 5.3 mmHg, 95% CI 0.6 to 10.0, p = 0.029), explaining approximately 29.8% (SBP) and 17.4% (DBP) of between-study heterogeneity.

##### Posture as moderator

Meta-regressions with posture (supine, seated, standing) did not detect effect modification for SBP (F = 1.1, p = 0.4; R² = 1.2%) or DBP (F = 0.9, p = 0.4; R² = 0.6%).

##### Risk of bias (overall) as moderator

No evidence of effect modification comparing Some/High vs Low risk: SBP β = −3.5 mmHg (95% CI −16.5 to 9.6, p = 0.582; R² ≈ 0%) and DBP β = −2.9 mmHg (95% CI −10.3 to 4.5, p = 0.417; R² ≈ 0%).

##### Dose–response and continuous modifiers (1-stage restricted cubic splines; REML with Hartung–Knapp)

Potassium supplementation was associated with clinically relevant reductions at moderate exposures. For assigned dose, the predicted change at 60 mmol/day was −8.8 mmHg for SBP (95% CI −14.8 to −3.1) and −4.8 mmHg for DBP (95% CI −8.0 to −1.7) **(Figure 4)**. For achieved urinary potassium (ΔK), the predicted change was −10.2 mmHg at 40 mmol/day (95% CI −18.4 to −2.1) and −5.6 mmHg at 60 mmol/day (95% CI −10.0 to −1.2) **(Figure 5)**. There was no strong evidence of nonlinearity across observed ranges (likelihood-ratio p ≈ 0.58–0.93), residual heterogeneity was modest (τ² ∼ 19.3–21.3 mmHg²). Longer duration predicted larger reductions (SBP β = −1.6 mmHg/week, SE 0.4, *p* < 0.001; DBP β = −0.8 mmHg/week, SE 0.2, *p* = 0.002), explaining ∼49% (SBP) and 35% (DBP) of heterogeneity. Higher baseline BP also predicted greater benefit (per +10 mmHg: SBP β = −4.1 mmHg, SE 1.2, *p* = 0.003; DBP β = −2.8 mmHg, SE 1.2, *p* = 0.03), with R² ≈ 43% and 22%, respectively. Assigned dose, ΔK and the K:Na ratio were not independently associated with effect size in univariable models (all *p* ≥ 0.1; R² ≤ ∼8%). Baseline urinary sodium showed a DBP-specific association (β = −0.8 mmHg per 10 mmol/day, SE 0.3, *p* = 0.03; R² ≈ 25%).

**Figure 4.**
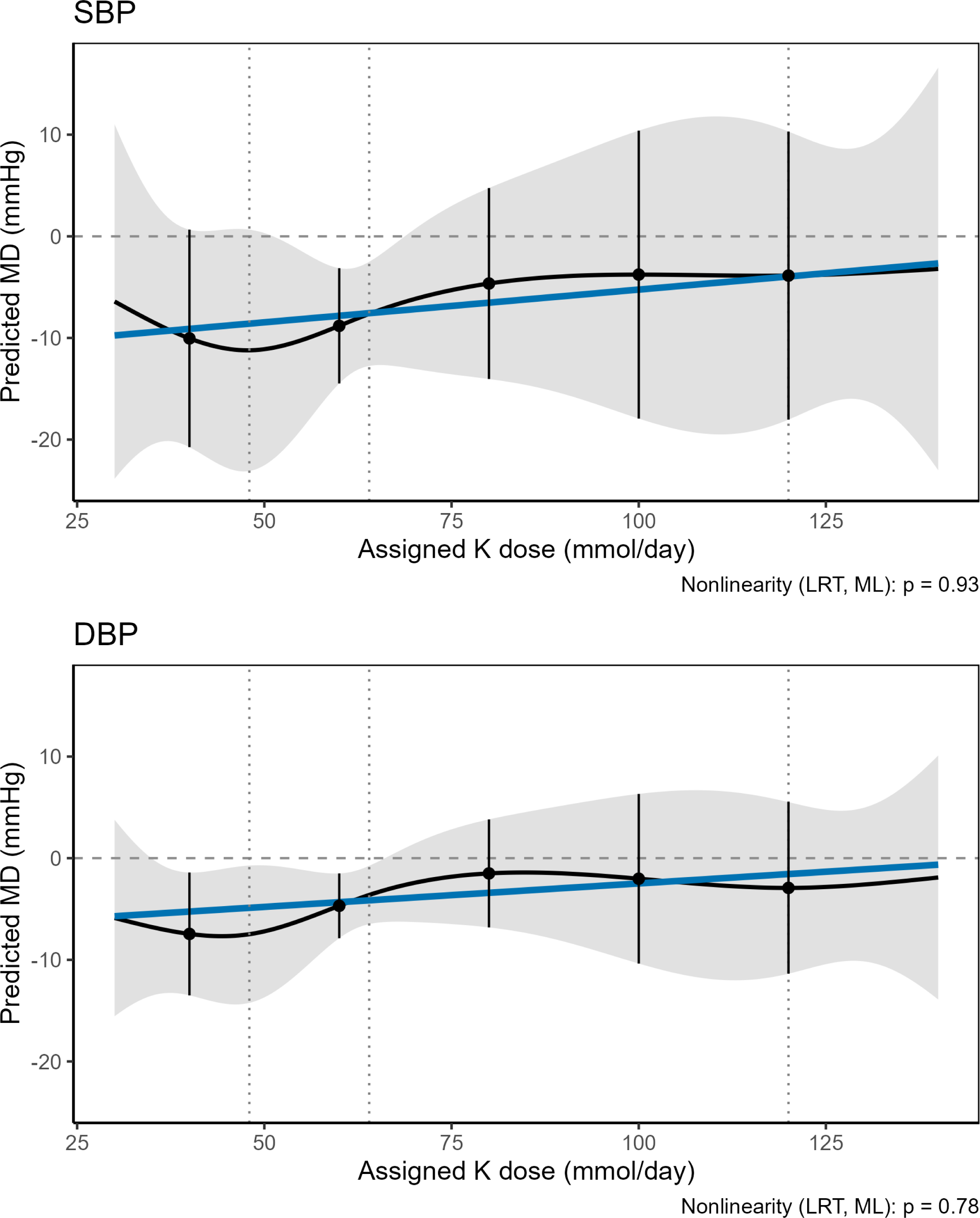
Dose–response relationship between assigned potassium dose and change in blood pressure. The black curve represents the fitted spline with 95% confidence bands (gray shading), while the blue line indicates the fitted *linear* meta-regression model. Vertical dotted lines mark knot locations.

**Figure 5.**
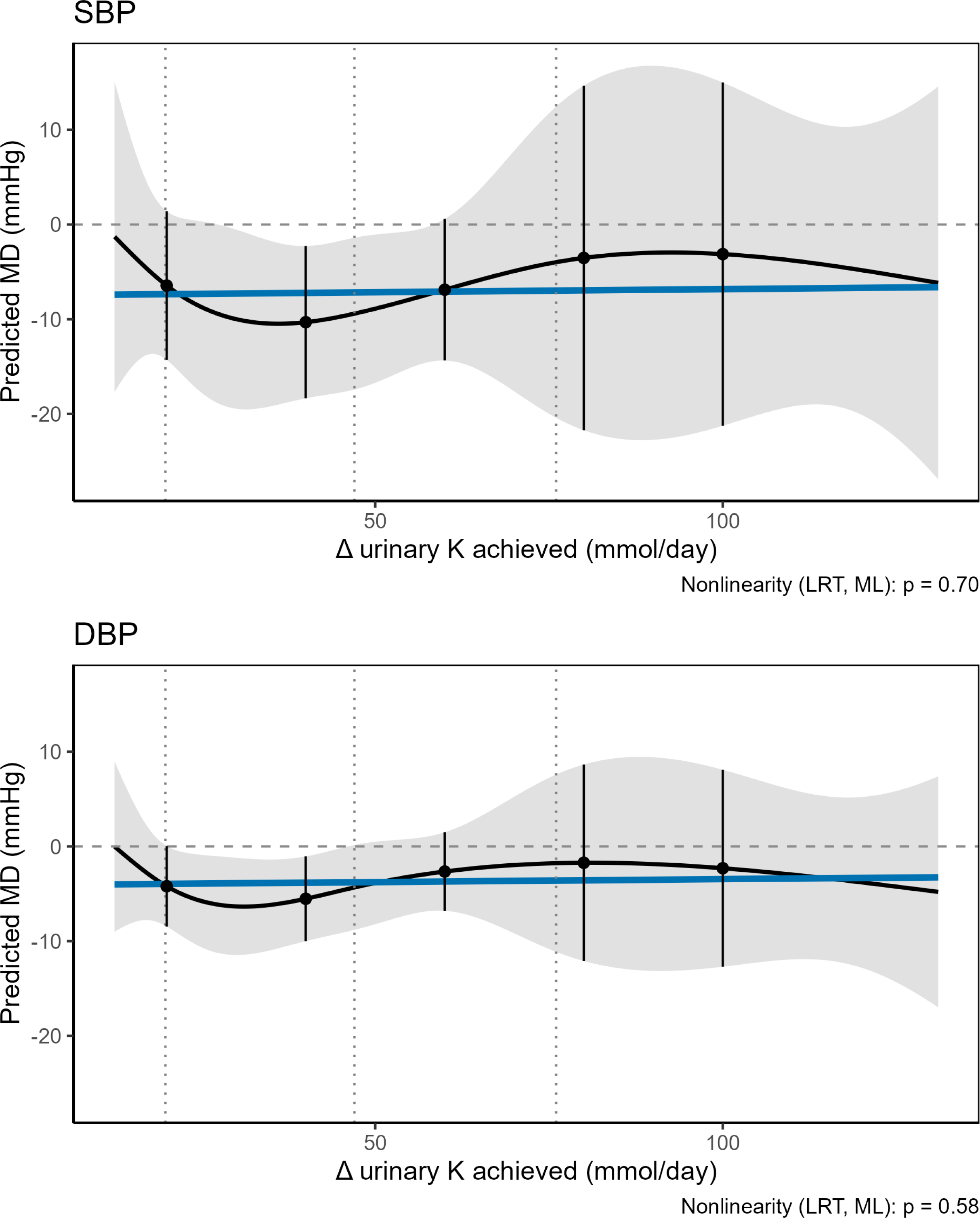
Dose–response relationship using achieved urinary potassium (ΔK) as exposure. The black curve represents the spline fit with 95% confidence bands (gray shading), and the blue line corresponds to the fitted *linear* model. Vertical dotted lines indicate knot positions.

##### Post-hoc multivariable meta-regression

We adjusted for study design (parallel vs crossover), trial duration, and baseline blood pressure for both SBP and DBP; for DBP we additionally included baseline urinary sodium and the ΔK:Na ratio. These covariates were chosen on methodological and biological grounds—design can affect precision and carryover, longer duration allows physiological stabilization, and higher baseline BP predicts larger absolute decreases—and because they showed signal in univariable screening with FDR control while preserving parsimony and adequate k. The models explained a large share of between-study heterogeneity (R²_het ≈ 77% for SBP; 76% for DBP) and yielded adjusted pooled effects of −7.5 mmHg for SBP (95% CI −10.3 to −4.6; PI −16.2 to 1.3) and −3.0 mmHg for DBP (95% CI −4.7 to −1.3; PI −7.6 to 1.6), indicating that differences in design, exposure time, and baseline electrolyte/pressure status account for much of the residual variance. Full outputs of all meta-regressions are presented in **Supplementary Table S9.**

Probability curves suggested the highest chances of clinically meaningful responses around 40–60 mmol/day (SBP ≤ −5 mmHg ≈ 65–70%; DBP ≤ −5 mmHg ≈ 60–65%; SBP ≤ −10 mmHg ≈ 40–45%; DBP ≤ −10 mmHg ≈ 20–25%). Full model outputs are provided in **Supplementary Table S10-S11** and **Figures S6–S7**.

### 20d. Results of syntheses (sensitivity analyses)

#### Fixed-effect sensitivity: clinic BP

Re-estimating the primary clinic BP meta-analyses with fixed-effect models yielded a common-effect MD of −5.33 mmHg (95% CI −6.25 to −4.40; k = 21) for SBP with very high heterogeneity (I² ≈ 92.3%). For DBP, the common-effect MD was −4.51 mmHg (95% CI −5.08 to −3.93; k = 21; I² ≈ 94.8%). These estimates are directionally consistent with the random-effects results and do not alter the clinical interpretation; given the substantial heterogeneity, random-effects remain the primary inference.

#### Fixed-effect sensitivity: ABPM

Using fixed-effect models for the ABPM analyses, the common-effect MD for SBP was −2.8 mmHg (95% CI −4.3 to −1.3; k = 7; I² ≈ 55%). For DBP, the common-effect MD was −3.2 mmHg (95% CI −4.2 to −2.1; k = 7; I² ≈ 84%). Results were directionally consistent with the random-effects analyses.

#### Measurement-error–corrected meta-regression (Δ urinary K)

Using meta-regression on the achieved change in urinary potassium per 10 mmol/day with SIMEX correction (random-effects REML with Hartung–Knapp), the slope for SBP was 0.02 mmHg (95% CI −0.20 to 0.24; p = 0.88; k = 18; I² ≈ 68%) and for DBP was 0.07 mmHg (95% CI −0.12 to 0.27; p = 0.47; k = 18; I² ≈ 88%). Estimates were virtually identical to uncorrected models (β ≈ 0.02 for SBP; 0.07 for DBP), indicating that measurement error in ΔK does not explain the lack of a linear association.

#### Risk-of-bias sensitivity (excluding high risk)

After excluding trials at high risk of bias, potassium supplementation remained associated with lower BP. For SBP, the pooled MD was −7.74 mmHg (95% CI −13.17 to −2.32; k = 15; I² = 93.7%) with a PI of −28.77 to 13.28. For DBP, the pooled MD was −4.04 mmHg (95% CI −7.07 to −1.01; k = 15; I² = 95.5%) with a PI of −15.77 to 7.69.

#### Dose–response (1-stage restricted cubic splines; four knots)

We found no evidence of departure from linearity for the association between potassium and BP. For models using Δ urinary K, the nonlinearity test was not significant for SBP (LRT, ML p = 0.8) or DBP (p = 0.71). Results were similar when using the assigned dose (SBP p = 0.79; DBP p = 0.48). Fitted curves closely tracked the linear trends, with the largest reductions around moderate exposures and wider uncertainty at the extremes; overall, no consistent inverse dose–response was observed. The sensitivity spline plots are provided in **Supplementary Figures S8** (assigned dose) and **S9** (Δ urinary K achieved).

#### Influence and outlier sensitivity

A composite diagnostic panel (studentized residuals, Cook’s distance, DFFITS, hat values) flagged one influential comparison for SBP (Obel, 1989) and two for DBP (Obel, 1989; Patki, 1990). Leave-one-out analyses showed the pooled SBP effect changed by at most 2.04 mmHg when omitting Obel, with a median absolute change of 0.22 mmHg and 95.2% of leave-one-out estimates within 0.5 mmHg of the full-model estimate. For DBP, the maximum change was 0.76 mmHg (at Obel), the median change was 0.16 mmHg and 90.5% of leave-one-out estimates were within 0.5 mmHg. Excluding the flagged SBP outlier reduced between-study heterogeneity substantially (τ² 65.14 to 9.86; I² 92.3% to 66.3%) while preserving the direction and significance of the effect (MD −5.21 mmHg; 95% CI −7.07 to −3.35; k = 20; PI95% −12.07 to 1.66). For DBP, excluding the flagged comparisons produced minimal change, consistent with negligible influence in the leave-one-out diagnostics.

#### Baujat analysis

Baujat plots indicated a single dominant contributor to heterogeneity and influence for SBP: Obel (1989), with the largest squared Pearson residual and greatest impact on the pooled estimate. For DBP, Obel (1989) again showed the highest influence and Patki (1990) moderate influence; all other comparisons clustered near the origin. These patterns align with the other diagnostics and the leave-one-out stability. The Baujat plot is presented as **Supplementary Figure S10**.

### 21. Reporting biases

We assessed small-study and publication biases only for meta-analyses with ≥10 studies, using visual inspection of funnel plots and the two-sided Egger regression test at α=0.10 within a random-effects framework. For outcomes with <10 studies, we did not conduct formal tests and provide only a narrative appraisal of the funnel.

For the office-based blood pressure outcomes, the Egger test was not significant for systolic blood pressure (SBP: p=0.53) or diastolic blood pressure (DBP: p=0.91). As a sensitivity analysis, we applied Duval and Tweedie’s trim-and-fill without replacing the primary estimates. For SBP, no studies were imputed (k₀=0) and the pooled effect was unchanged (random-effects mean difference [MD] before = −7.199 mmHg; after = −7.199 mmHg). For DBP, four studies were imputed (k₀=4), shifting the pooled MD from −3.954 to −4.966 mmHg (**Supplementary Figures 11-12**). These trim-and-fill results are interpreted cautiously, as the procedure does not establish publication bias and can be influenced by between-study heterogeneity.

For the ABPM outcome, fewer than ten studies were available; therefore, no formal assessment of small-study or publication bias was performed. Visual inspection suggested no obvious asymmetry, but this should be interpreted with caution due to the small number of comparisons.

For any adverse event (k = 13) and total withdrawals (k = 12), no asymmetry was detected (Egger p = 0.53 and 0.69, respectively). In contrast, for withdrawals due to adverse events (k = 17), the funnel plot showed marked asymmetry (p < 0.001), suggesting the presence of small-study effects or selective non-reporting. Trim-and-fill analyses did not converge, therefore, no imputed studies were added. Overall, publication bias cannot be excluded for the adverse-event withdrawal outcome.

#### Risk of bias due to missing evidence (ROB-ME)

We applied the ROB-ME tool to each meta-analysis to evaluate bias arising from non-reporting within studies and from potentially missing studies across the review. The historical nature of this evidence base (several trials pre-registration) indicates a plausible risk that study identification depended on results. The full ROB-ME assessment is provided in the Supplementary Materials.

##### Efficacy outcomes (MA-1, MA-2, MA-3, MA-4)

For clinic SBP (MA-1) and DBP (MA-2), all eligible trials reported end-of-treatment values; within-study non-reporting was not suspected. Across-study concerns remain because many trials pre-date routine registration; however, small-study tests were not significant and trim-and-fill suggested at most modest shifts. ROB-ME judgments were Some concerns. The predicted direction of any residual bias was away from the null for SBP (undetected null studies would attenuate benefit) and towards the null for DBP (any bias would make the observed effect conservative). For ABPM 24-h SBP/DBP (MA-3/MA-4), k<10 precluded formal tests; with sparse data and mixed reporting historically, ROB-ME was Some concerns, with unpredictable direction.

##### Harms and withdrawals (MA-5, MA-6, MA-7)

For withdrawals due to adverse events (MA-5), reporting was heterogeneous and the funnel plot showed marked asymmetry; trim-and-fill did not converge. ROB-ME judgment was High risk of bias, with predicted direction away from the null (i.e., the current RR is likely inflated and would attenuate with complete evidence). For any adverse event (MA-6), omissions in older trials reflect generic incompleteness rather than selective non-reporting; small-study tests were not significant. ROB-ME judgment: Some concerns, predicted direction away from the null (if unreported results were more often null). For total withdrawals (MA-7), only one eligible trial had unclear extractability; small-study testing showed no asymmetry. ROB-ME judgment: Some concerns, predicted direction away from the null.

### 22. Certainty of evidence (GRADE)

Oral potassium supplementation reduced clinic blood pressure by a modest–moderate amount (SBP mean difference [MD] −7.2 mmHg, 95% CI −11.1 to −3.4; DBP MD −4.0 mmHg, −6.2 to −1.7; low certainty). For 24-hour ambulatory BP (ABPM) the effects were smaller and very uncertain (SBP MD −2.4 mmHg, −5.2 to 0.4; DBP MD −1.6 mmHg, −4.2 to 1.1; very low certainty). Any adverse event was more frequent (risk ratio [RR] 2.41, 1.63–3.56; high certainty), which corresponds to 40 more per 1,000 participants (17–75 more). Withdrawals due to adverse events probably increased slightly (RR 1.28, 1.04–1.59; low certainty; 0–10 more per 1,000). Total withdrawals also probably increased slightly (RR 1.32, 1.03–1.70; low certainty; 4 more per 1,000, 0–9 more). Summary of Findings is provided in the **Supplementary Materials**.

#### Recommendation

Conditional recommendation for the intervention. We suggest oral potassium supplementation, in addition to usual care, for adults with untreated primary hypertension provided renal function is normal and basic laboratory monitoring is available.

#### Rationale

Average clinic BP reductions are modest, effects on ABPM are very uncertain. Harms increase in absolute terms by small amounts. Given low cost and good feasibility with routine labs, the overall balance of effects probably favors offering supplementation to selected patients rather than as a blanket policy.

## DISCUSSION

### 23a. Discussion (interpretation)

#### Clinical efficacy versus ambulatory efficacy

We observed mean reductions in office blood pressure measurements (−7.2/−4.0 mmHg for SBP/DBP) accompanied by high heterogeneity, and modest or nonsignificant decreases in ABPM (−2.4/−1.6 mmHg). This pattern is consistent with a *Journal of Hypertension* review comparing treatment-induced responses between office and ambulatory readings, which showed that the 24-hour mean reduction is systematically smaller than that observed in clinic measurements (for both SBP and DBP), and smaller at night than during the day for DBP, with an average ABPM/office ratio of 0.67–0.75 (20). In our meta-analysis, this ratio was lower (0.33 for SBP and 0.40 for DBP), likely influenced by the limited number of trials reporting ABPM (k=6), which reduces precision and may underestimate the 24-hour effect. Furthermore, clinical evidence indicates that changes measured by ABPM and office readings are not in a 1:1 correspondence, since ABPM attenuates the white-coat effect and identifies pseudoresistance (patients apparently uncontrolled in clinic but normal over 24 h) as well as masked hypertension. Consequently, treatment-induced reductions observed through ABPM tend to be smaller than those in the office (21).

For office blood pressure, the mean reduction observed is directionally consistent with prior evidence on potassium supplementation or increased dietary potassium intake. For instance, Aburto et al. reported reductions of −3.49 mmHg for SBP and −1.96 mmHg for DBP, while Poorolajal et al. found −4.25 mmHg and −2.53 mmHg, respectively (22,23). In our analysis, the effect remained across all sensitivity checks: using a fixed-effects model, SBP −5.33 and DBP −4.51; when restricted to trials with lower risk of bias, SBP −7.74 and DBP −4.04; and after excluding the most influential study, SBP −5.21 with heterogeneity (I²) dropping from 92.3% to 66.3%, while DBP changes were minimal. These findings suggest that the office-based effect is robust and not driven by any single study or analytic decision.

#### Prediction intervals

Unlike previous reviews on potassium and blood pressure—which report only mean effects with 95% CIs—our study incorporates explicit prediction intervals, allowing a translational reading of the range expected in a future trial. In our results, the office SBP had a PI of −24.5 to +10.1, and DBP −14.1 to +6.1. For ABPM, PIs were SBP −8.8 to +3.9 and DBP −8.8 to +5.7.

These intervals show that, even with a favorable overall mean, there are plausible contexts where the reduction may be small or absent; hence the advisability of avoiding universal recommendations, calibrating expectations, and confirming responses with out-of-office measurements whenever possible. This between-context variability is further explored below through the analysis of effect modifiers.

#### Effect modifiers and explanation of heterogeneity

The observed patterns are congruent with the physiology of sodium–potassium balance and with comparative literature. The greater reduction in participants with higher baseline BP is compatible with a larger hemodynamic margin and higher sodium sensitivity; in our data this translated into additional SBP decreases of about 4 mmHg for every 10 mmHg increase in baseline BP. Likewise, longer interventions showed larger reductions, consistent with the time needed to reach sodium–water balance steady state and consolidate vascular changes; we estimated an additional SBP decrease of about 1.6 mmHg per week of exposure.

The difference between parallel and crossover trials also has a clear clinical–methodological basis. In crossover designs, washout periods are often insufficient to fully reverse renal and vascular potassium effects, and protocol learning may attenuate intra-individual variability, masking modest changes. In contrast, parallel trials better capture cumulative effects and minimize carryover between periods, which was reflected in larger pooled estimates in our synthesis.

These observations align with previous meta-analyses focused on total potassium intake or urinary excretion, which described more pronounced benefits in hypertensive subjects and with adequate exposure, but without substantively quantifying between-study variability (22–25). In our case, a multivariable meta-regression integrating design, duration, and baseline BP explained roughly 75–77% of heterogeneity while maintaining clinically relevant adjusted effects (SBP = −7.5 mmHg; DBP = −3.0 mmHg). For DBP, baseline urinary sodium and the K:Na ratio emerged as relevant covariates: baseline sodium was associated with greater reductions in univariable analysis, and, together with K:Na, improved the multivariable model fit. The signal was clearer for DBP than for SBP, which is physiologically plausible: DBP primarily reflects peripheral vascular resistance—the component most sensitive to the combination of natriuresis and endothelium-dependent vasodilation induced by potassium—whereas SBP, more influenced by large artery stiffness and stroke volume, shows less modulation over the timeframes of these trials and greater measurement variability, so the contribution of baseline sodium tends to attenuate after adjustment for baseline BP and duration. This pattern is consistent with known physiology: potassium increases promote natriuresis (via NCC inhibition in the distal tubule) (26–28), modulate the renin–angiotensin–aldosterone system (29,30) and induce endothelium-dependent vasodilation (31,32); these effects are more pronounced when the initial Na⁺/K⁺ gradient is displaced toward sodium excess (33).

In summary, the blood pressure–lowering effect of potassium appears to act primarily through restoration of the ionic balance disrupted by sodium, which also helps explain the breadth of prediction intervals and supports a conditional recommendation tailored to clinical context. Given its post hoc nature and the limited number of studies per covariate, we interpret these findings as explanatory and hypothesis-generating, guiding candidate selection (higher baseline BP, sustained exposure) and the design of future trials (parallel, adequate duration).

#### Dose–response relationship

In our dose–response models, neither the assigned dose nor the 24-hour urinary ΔK showed a clear association with BP change; tests for nonlinearity were nonsignificant and no robust thresholds were identified. This contrasts with Filippini et al. (24), who described a U-shaped curve when exposure was measured as the between-arm difference in urinary excretion: the benefit weakened above 30 mmol/d and, with differences ≥80 mmol/d, BP increased; the effect was greater in hypertensives and with high sodium intake (noting few trials at the high end and predominance of crossover designs).

Using the same spline framework, we did not observe a global dose–response curve, likely because our exposure range (dose/ΔK in an intermediate range) provides less information at the extremes. Our contribution is complementary within the dose–response spectrum: we show that, within the usual clinical range in hypertensive patients, the isolated dose explains little, and the response is modulated by sodium context (better fit for DBP when baseline sodium and K:Na are included), suggesting that the hypotensive effect arises mainly from reestablishing Na⁺/K⁺ balance rather than from simple dose escalation.

#### Patient-oriented safety

Beyond biomarkers, we synthesized safety outcomes in clinically meaningful terms. Across available RCTs, the likelihood of experiencing ≥1 adverse event (AE) was higher with supplementation (RR 2.41), corresponding to 40 additional AEs per 1,000 individuals (compatible range 17–75, NNH 25). Withdrawals due to AEs increased slightly (RR 1.28–1.30), with a very small absolute change: 0–10 additional withdrawals per 1,000. Total withdrawals also rose marginally (RR 1.32; 4 more per 1,000, interval 0–9). Reported AEs were mostly mild and gastrointestinal. The synthesized evidence does not allow consistent quantification of serious AEs due to heterogeneity and incomplete reporting. Overall, in adults with normal renal function and not taking potassium-elevating drugs, the safety profile appears reasonable under basic monitoring (renal function and serum K⁺ at baseline and 1–2 weeks or after dose/medication changes), with explicit communication of risk in shared decision-making.

### 23b. Discussion (limitations of evidence)

The evidence presents several limitations. First, the number of trials with ABPM is small, leading to imprecision and limited inference beyond office settings. Second, crossover designs predominate, and only five trials reported washout periods of 2–5 weeks, while the rest did not report or implement them; this increases carryover risk and may attenuate or bias effects. Third, trials were short (median 4 weeks), leaving effect durability and rare events uncertain. Fourth, we observed high heterogeneity in office BP, and ABPM effects were small and imprecise, limiting the applicability of a single mean estimate. Fifth, measurement and quantification of baseline/consumed sodium were not uniform across studies, hindering interpretation of ionic-context modulation. Sixth, for AE-related withdrawals, there were indications of small-study/publication bias, and GRADE certainty for ABPM outcomes was very low. Taken together, these aspects call for cautious interpretation of the benefit magnitude and its generalizability.

### 23c. Discussion (limitations of review processes)

Despite a comprehensive search, attempts to obtain missing data through author contact were unsuccessful, leaving some uncertainties unresolved. Study selection and data extraction were performed in duplicate, minimizing though not eliminating errors. For rare outcomes, we applied a continuity correction of 0.5; alternative decisions might slightly alter confidence intervals. Finally, the prespecified multivariate model to jointly analyze office and ambulatory BP could not be implemented due to insufficient studies, so outcomes were synthesized separately.

### 23d. Discussion (implications)

#### Practice and policy

The findings suggest a modest yet clinically relevant benefit in office BP, with greater uncertainty for ABPM. In practice, potassium supplementation may be conditionally considered for adults with primary hypertension and normal renal function, ideally combined with sodium reduction and other lifestyle measures. Individualization is essential: check creatinine and K⁺ at baseline and after initiation or dose changes; avoid or exercise caution in the presence of potassium-elevating drugs or chronic kidney disease. To assess response and avoid misclassification due to the “white-coat effect,” prioritize ABPM or home BP monitoring. At the policy level, the results support public health interventions focused on Na⁺/K⁺ balance, including nutritional education and, where appropriate, potassium supplementation within safety frameworks and risk-population surveillance.

#### Future research

Future RCTs should adopt parallel designs lasting ≥8–12 weeks, with prespecified ABPM, prospective registration, and paired reporting in crossovers (variances, adequate washouts). Priorities include: (1) dose–response and differentiation by salt form (e.g., KCl vs. citrate); (2) effect modifiers (baseline BP, sodium/potassium intake or excretion, K:Na ratio); (3) medium-term safety in subgroups (CKD, ACEI/ARB/potassium-sparing drugs) with standardized AE reporting; (4) pragmatic primary care trials combining potassium supplementation with sodium reduction and patient-centered outcomes; (5) individual participant data meta-analyses to refine candidate selection and expected effect magnitude. This agenda will enable progression from a conditional recommendation to more precise guidance by patient profile and ionic context.

## CONCLUSIONS

Oral potassium supplementation produces modest, clinically relevant reductions in clinic blood pressure in adults with untreated primary hypertension, while effects on 24-hour ambulatory blood pressure are smaller and very uncertain. Safety findings indicate a higher risk of experiencing at least one adverse event—predominantly mild, gastrointestinal—with only small absolute increases in withdrawals. Taken together, these results support a conditional use of potassium supplementation alongside lifestyle measures—especially sodium reduction—in carefully selected patients with normal renal function and basic laboratory monitoring. Given the wide prediction intervals and heterogeneity partly explained by baseline pressure, duration, and sodium–potassium context, decisions should prioritize out-of-office monitoring (ABPM or home BP) to verify response. Further adequately powered parallel-group trials of ≥8–12 weeks with prespecified ABPM and standardized safety reporting are warranted to refine candidate selection and expected benefit.

## Data Availability

Data used for all analyses will be made available upon reasonable request from the corresponding author. Requests will be reviewed to ensure appropriate use and compliance with ethical and institutional requirements.

## OTHER INFORMATION

### 24a. Registration and protocol (registration)

This review was prospectively registered in PROSPERO (ID: CRD42024621287); the record is published and stable.

### 24b. Registration and protocol (protocol)

The full protocol is publicly available in the PROSPERO record. Any amendments made after registration are documented in the registry and described in item 24c.

### 24c. Registration and protocol (amendments)

During the data extraction phase (after registration and study selection but before quantitative synthesis), we introduced methodological clarifications to enhance clarity, transparency, and alignment with current standards. We defined clinic blood pressure and ambulatory blood pressure (ABPM) as co-primary outcomes; clarified the handling of continuous outcomes (final values as the default metric and direct derivation of standard errors from mean differences with confidence intervals where applicable); specified urinary potassium and sodium as exposure/adherence indicators rather than outcomes; refocused subgroup analyses on clinically relevant modifiers; added ROB-ME to assess bias due to missing evidence; and pre-specified decision-oriented visualizations (probability curves and benefit–risk plots). These changes did not alter eligibility criteria or the set of included studies. The full, time-stamped version of these amendments is available in PROSPERO.

### 25. Support

This review received no external funding and was self-funded by the authors. We acknowledge Universidad Peruana Cayetano Heredia (UPCH) for non-financial support in the form of methodological guidance from an academic advisor. Apart from this advisory support, neither UPCH nor any other funder, sponsor, or institution had any role in the study design, protocol development, study selection, data collection/management, analysis, interpretation, writing, or the decision to submit or publish; all decisions were made by the authors.

### 27. Availability of data, code, and other materials

Data used for all analyses will be made available upon reasonable request from the corresponding author (Javier Ravichagua). Requests will be reviewed to ensure appropriate use and compliance with ethical and institutional requirements.

